# Integrating Community-Based Health Information System with a Patient-Centered Medical Home to Improve Care of Patients with Hypertension: A Longitudinal Observational Study Protocol

**DOI:** 10.1101/2023.07.09.23292420

**Authors:** Unab I. Khan, Sabeen Shah, Shankar Viswanathan, Asra Qureshi, Yasmeen Noornabi, Mahnoor Niaz, Judith Wylie-Rosett

**Author notes:** Corresponding Author: Unab I. Khan MBBS, MS., Associate Professor, Department of Family Medicine, Aga Khan University, PO Box 3500, Stadium Road campus, Karachi 74800, Pakistan.

## Abstract

**Introduction:** Vertical health delivery models in Pakistan focus on providing episodic, disease-based care. Health care for middle-class communities is largely through a fee-for-service model that ignores preventive care. The growing burden of cardiovascular illnesses requires restructuring of primary health care system allowing well-coordinated efforts between patients and providers. We propose a model of care that integrates a Patient-Centered Medical Home (PCMH) with a Community-Based Health Information System (CBHIS) for bidirectional communication at the patient and community level. This protocol describes the integration and evaluation of the PCMH-CBHIS infrastructure using hypertension (HTN) as a model.

**Methods:** This is a population-based, observational, longitudinal study in an urban setting in Pakistan. Through convenience sampling, participants will be enrolled in CBHIS and followed longitudinally over two years for HTN outcomes. A mixed-methods approach will be used to evaluate the process of integrating PCMH with CBHIS. This will involve building partnerships with the community through formal and informal meetings, focus group discussions, and a household health assessment survey. Community members identified with HTN will be linked to PCMH for disease management and skills to improve self-management. A customized electronic medical record system will be developed to link community-level data with family and patient-level data to track changes in disease burden. The RE-AIM evaluation framework will be used to monitor community and individual-level metrics to guide implementation assessment, the potential for generalization, and the effectiveness of the PCMH in improving health outcomes.

**Ethics and dissemination plan:** Ethical clearance was obtained from Ethics Review Committee at Aga Khan University (2022-6723-20985). We plan to present the findings from this research at conferences and publish them in peer-reviewed journals. Additionally, we intend to leverage findings from this research to obtain funds focusing on chronic disease care in similar settings.

**Article Summary:** *Strengths and limitations of this study:*

- The study integrates a comprehensive health assessment survey to assess the disease burden at the community level to guide the prioritization of health services and prevention efforts at the health facility.
- A mixed methods approach will be used to measure the effectiveness of hypertension management through the proposed model of care.
- The development of a customized electronic medical record system will allow aligning clinic- and community-based activities.
- Community members may migrate from the catchment area, limiting the longitudinal assessment of hypertension management.
- With a patient-driven model, we cannot predict how many community participants would choose the health facility for ongoing care.

## INTRODUCTION

Pakistan, which has a population of nearly 240 million, faces a double burden of communicable and non-communicable diseases (NCDs) (1). Deaths due to NCDs have increased from 46% in 2000 to 60% in 2019 (2), and cardiovascular disease (CVD) (including ischemic heart disease and stroke) remains the leading cause of these deaths (3). Hypertension (HTN) is the leading modifiable CVD risk factor, and one in four adults is diagnosed with HTN (4). In addition, blood pressure (BP) control rates as low as 6.4% have been reported at the primary care level (5), contributing to more than half of cardiovascular disease-related morbidity (6).

The fragmented healthcare system in Pakistan relies heavily on the private sector, especially in urban areas, where 70% of care is delivered at private clinics and hospitals with a fee-for-service model (7). With rising inflation, health care is becoming increasingly expensive, leading to episodic, disease-specific care with little or no focus on health promotion and disease prevention. The financial burden of chronic disease management creates additional challenges to compliance, further contributing to morbidity and mortality. This is especially true for the middle class, who are largely ineligible for government-funded health programs and rely on unmonitored and unregulated private-sector healthcare (8).

Middle-class (MC) in Pakistan is defined as households with the ability to spend $2-10 per person/day, and low-middle class is defined as households with the ability to spend $2-4/person/day (9, 10). MC households have a secure income source, can spend beyond essential goods, and can pay for health and education. However, if struck by catastrophic health crises, the financial implications of illness can plunge an MC family below the poverty line (11). In 2011, MC constituted 55% of households, with an estimated 84 million people (12); and the average growth between 1999-2019 is 9.4% per year (9).

The Aga Khan University’s health research mandate is to improve socio-economic determinants of health. In this vein, the University has funded a primary care center, Family Medicine Health Center (FMHC) to create a proof-of-concept health model that, if effective, can be replicated by other health systems. Using the principles of a Patient-Centered Medical Home (PCMH), FMHC provides accessible, cost-effective, quality care in the community setting (13-15). In addition, it integrates with a Community-Based Health Information System (CBHIS), to examine the effectiveness of the model on health outcomes at the patient and community level.

As there is no national integrated health information system that collects health parameters, we are unable to make data-driven decisions at the population level (16). Thus establishing a CBHIS for continuous monitoring will be essential (17, 18). We will create a CBHIS for the catchment population around FMHC to measure the disease burden and create baseline estimates that will enable us to assess change at the individual, family, and community levels.

Figure 1 denotes the juxtaposition of the CBHIS with a PCMH model at FMHC, illustrating the steps we will undertake to create a system of ongoing bilateral communication between the community and the FMHC team. A series of dialogues between the community and the FMHC team will be organized for the formation of a Community Advisory Board (CAB), that will support planning and conducting focus group discussions (FGD) with community members and initiate a family-level health assessment survey (HAS) for all families within a 0.5-kilometer radius of FMHC. Information from the initial HAS and FGDs will form the basis of the CBHIS, and annual surveys will be conducted to examine the impact of FMHC-based interventions at the community level. In addition, subsequent FGDs will allow us to understand the context of the responses in HAS. We will use an electronic medical record (EMR) system to connect the patient-level data from FMHC to community-level data in CBHIS. The current manuscript describes the protocol for integrating and evaluating the PCMH-CBHIS infrastructure to improve the health outcomes of patients diagnosed with HTN within the catchment area of FMHC.

**Figure 1.**
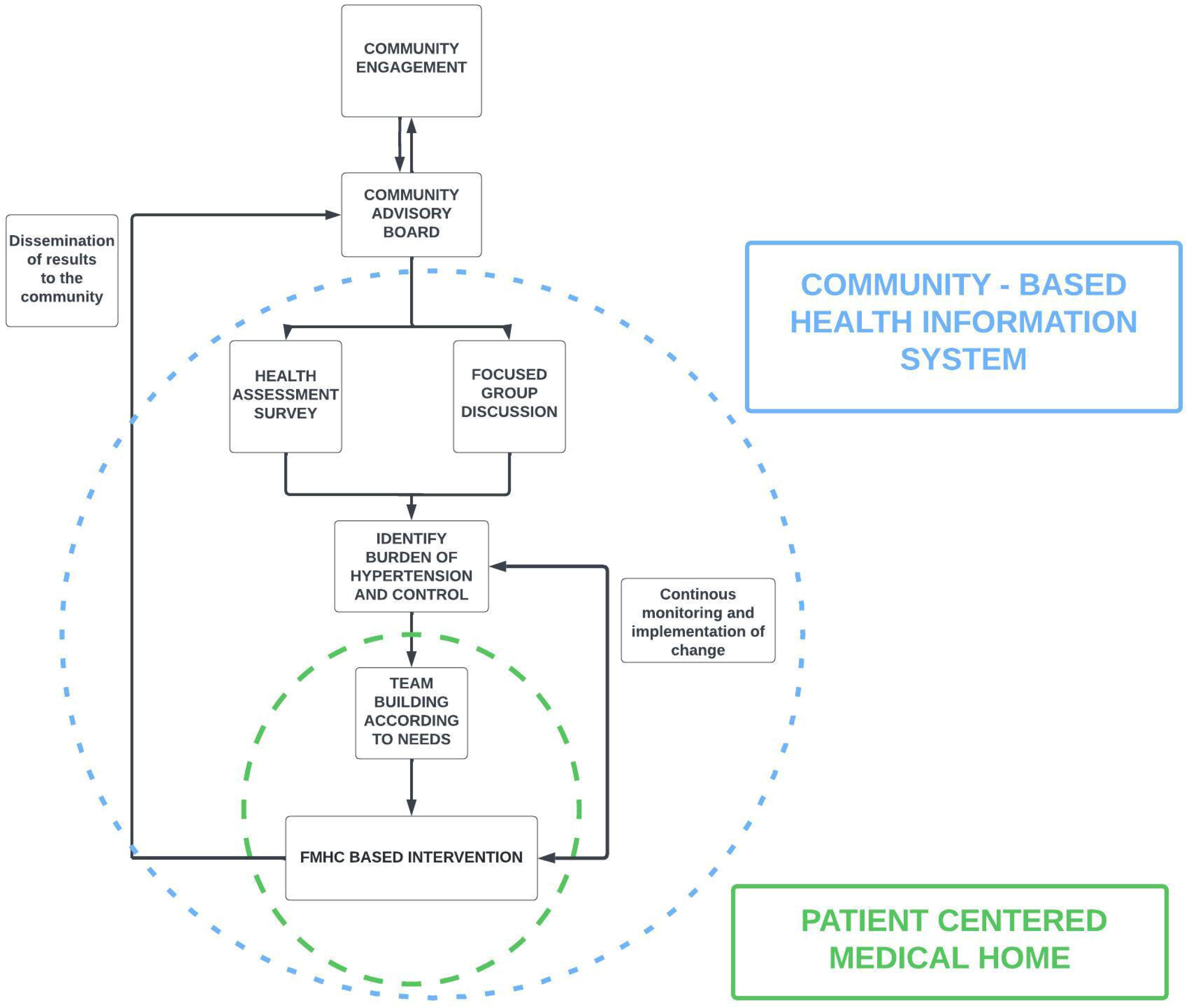
Steps Towards Integrating a Community-Based Health Information System with a Patient-Centered Medical Home Model

## MATERIALS AND METHODS

### Study setting

FMHC is located in District Central of Karachi. The district’s area is 69 km^2^ with a population density of 43,063.51 persons per km^2^ (19, 20). Most people live in apartment complexes. Our catchment population includes families residing within a 0.5-kilometer (km) radius of FMHC.

### Study design

This is a population-based, observational, longitudinal study. Participants will be enrolled in CBHIS through convenience sampling of the catchment population. We will use a mixed-methods approach to evaluate the process of integrating PCMH with CBHIS.

### Study period

The preliminary implementation and evaluation will be conducted over 24 months between June 2022 to June 2024.

### Sample estimates

Based on 2017 census, the population of District Central is 2,971,382 (20). With an estimated 45% of the total Pakistani population reported to be children ≤ 18years of age (21), we estimate that 1,634,260 adults (>18 years) are residing in District Central with a population density of 23,685 adults/km^2^ (22). We have defined the catchment population for FMHC to be people living within a 0.5 km radius of FMHC, which is estimated to have 37,185 adults. Considering the prevalence of HTN in urban Pakistan as 33% (21), we assume that an estimated 12,270 adults in the catchment area will have HTN. We expect to serve 20% of patients with HTN at FMCH, thus aiming to improve health outcomes for an estimated 2,454 adults with HTN.

### Creation of a Community-Based Health Information System

A multidisciplinary team-based approach will be adopted to create and maintain the CBHIS, with community involvement vital to optimizing and implementing ongoing activities.

### Community Engagement

#### Formal and informal meetings

We will introduce the creation of a CBHIS and its utility in health provision at FMHC to community leaders, faith leaders, business leaders, and other key stakeholders in the catchment area. These meetings will also allow us to understand the community’s cultural dynamics, current options of care, their openness to try alternative options, and identify ways of ensuring maximal community participation. Our goal is the formation of a CAB that can help with ongoing bidirectional communication, ensuring participation by families in creating and maintaining the CBHIS and informing them about services at FMHC. As stakeholders, CAB members will also provide inputs into study methods, such as the feasibility of hiring community members for conducting the HAS and identifying volunteer families for piloting HAS.

#### Focus group discussions

To better understand the community’s health-related needs, we will conduct exploratory FGDs in the catchment area. FGDs will continue till we reach data saturation in the responses. Preliminary focus groups will explore: 1) the community’s perception of quality health care, 2) self-perception of health needs, 3) self-perception of health status, 4) barriers to access care, 5) attributes of a facility perceived to offer quality care and 6) the willingness and paying power for quality care. We will work with community leaders to provide a private space accommodating 15-20 people where possible. Respecting cultural nuances, we will conduct separate FGDs for men, women, youth, and key stakeholders. Subsequent FGDs will be conducted amongst a representative sample for insights on FMHC-based interventions (such as barriers to accessing care at FMHC).

#### Analysis of focus group discussions

Audiotapes of FGDs will be transcribed in Urdu (local language), and then translated into English. We will verify the translated discussion against the audiotapes for accuracy. English version of the transcript will be prepared in text files and analyzed manually. Three members of the team will complete the coding independently. We will then obtain a consensus on the correlation between codes and themes.

### Health Assessment Survey (HAS)

The HAS will form the basis of the CBHIS, which will serve as a platform to understand the disease burden of the community and guide in prioritizing service delivery and prevention efforts at FMHC. Survey questions will be derived from existing national standardized surveys and modified to cultural context (22-24). We will conduct a door-to-door survey of all consenting families in the catchment area at baseline. We will consider each apartment as a household with one family, using the definition “Person or group of related and unrelated persons who live together in the same dwelling unit(s) or in connected premises, who acknowledge one adult member as head of the household, and who have common arrangements for cooking and eating” (23). However, if there is subletting, or the extended family has separate income and expenses, each will be considered an independent family within the apartment. The survey will include information at the family level (demographics, family income, healthcare expenditure) as well as individual-level data (such as medical history, healthcare utilization, anthropometrics (height, weight, waist circumference), BP, and perception of health). Subsequent annual surveys will be conducted to evaluate changing demographics and disease trends.

#### Survey administration

The survey will be conducted in Urdu, the local language. We will pilot test HAS in the community to determine face validity. Piloted and ERC-approved versions of HAS will be incorporated as an e-questionnaire, and data will be stored in REDCap (25). Trained community health workers (CHWs) will directly record the information on Android tablets. After informed consent, the survey will begin with the head of the family, followed by all other consenting members. To ensure representation of both sexes, CHWs will approach the male community members at the local mosques following congregational prayers, whereas female community members will be approached at their homes after school hours.

#### Selection and role of community health workers

With input from CAB members, we will recruit CHWs (from within or outside the catchment area) who will, 1) map the catchment area; 2) conduct surveys; 3) strengthen connections between the community and FMHC team through care coordination, such as by helping community members with HTN access FMHC services; and 4) provide informal counseling on healthy behaviors.

#### Training of community health workers

A two-day training will be conducted to enhance CHWs’ competency in 1) soft skills such as respectful communication with peers and community members, time management, etc.; 2) technical skills such as the use of REDCap for HAS administration; 3) clinical skills to obtain blood pressure (BP) and anthropometric measurements; 4) knowledge about evidence-based algorithms modified for community settings to facilitate prompt identification of uncontrolled BP and early linkage to FMHC. The research team will monitor data for accuracy, completeness, and consistency every week and provide feedback to the CHWs.

#### Data Analysis of HAS

We will use descriptive statistics to understand 1) the burden of HTN and other non-communicable diseases in the community; 2) HTN control rates; 3) the distribution of CVD risk factors (such as smoking, obesity, and physical activity); and 4) health care utilization (outpatient and hospitals settings). The prevalence and Clopper-Pearson exact 95% confidence interval will be presented. Additionally, sub-group analyses will be performed using appropriate statistical tests (e.g., ANOVA, Chi-square) comparing demographics, health expenditure, access to care, and presence of comorbid illnesses among patients with 1) controlled and uncontrolled HTN; and 2) patients at FMHC and FMHC non-users.

### Optimization of Services at Family Medicine Health Center

The FMHC team will offer evidence-based clinical care and use Shared Decision Making (26) and onsite educational sessions to deliver patient-centered care and improve the health literacy of FMHC patients with HTN (26). These interactions will incorporate environmental and social determinants of health to stimulate self-management in HTN through a heart-healthy diet, reduction in sodium intake, increased physical activity, reduction in overweight and obesity, as well as improved knowledge of HTN risk factors. The on-site, small-group educational sessions will be publicized to the wider community by CHWs and CAB members. Additionally, CHWs will invite CBHIS participants identified with elevated BP or HTN over the phone or in person during routine field visits (27).

If needed, specialty referrals will be arranged at AKU’s community hospital in Karimabad which is 1.5 km from FMHC.

The research team (CHWs, statisticians, and public health specialists) and the clinical team at FMHC will meet regularly to share insights about community-level and patient-level issues.

Training for the clinical team: A two-day training of FMHC doctors and nurses will be conducted to enhance clinical knowledge about HTN management and CVD risk assessment based on the WHO recommendations (28). This will include various aspects of HTN care to administer age – and sex-specific care packages in an opportunistic manner that support: 1) diagnosis of HTN; 2) CVD risk assessment and stratification; 3) treatment initiation using appropriate antihypertensive drug class; 4) counseling about lifestyle change, modified to cultural context; 5) medication dose adjustment; 6) recognition of red flags and points of referral; and 7) introduction to the monitoring and evaluation indicators.

The scope of training for nurses will cover 1) standardized BP and weight measurement, 2) diagnosis of HTN, 3) counseling about lifestyle change, modified to cultural context, 4) early recognition of red flags and points of referral, and 5) introduction to the monitoring and evaluation indicators. Ongoing capacity-building sessions will be geared towards refresher training and the inclusion of new knowledge and skills to meet evolving needs of FMHC services.

### Health Information Technology

FMHC has a customized electronic medical record (EMR) that uses ICD-11 codes for standardized data entry (29). This maximizes the potential of EMR to provide structured data for analyzing changes in disease trends and recognizing upcoming health issues at individual, family, and community levels. The EMR can incorporate the individual patient’s HAS data from CBHIS into their medical record using a unique survey number. This will allow the healthcare team access to survey data (including barriers to care and household expenditure) guiding personalized counseling based on identified barriers to care to enhance medication adherence and self-efficacy in HTN management. Additionally, to support the healthcare team in providing cost-effective care EMR will include the cost of different brands of antihypertensive medications to allow physicians to identify affordable medications based on patients’ paying power (30). Furthermore, the EMR interface has been designed to improve the overall quality and efficiency of the FMHC team by strategically placing information about a single patient on one screen enabling a quick overview of all necessary details.

#### Analysis of EMR data

A dashboard will provide access to data in real time to facilitate interpretation and ensure the availability of information for auditing and quality improvement. Patient-level data will be analyzed to follow trends of HTN control and how this varies with other health parameters such as body mass index (BMI) and biochemical measurements (such as serum creatinine level, cholesterol level, and fasting glucose level). Additionally, EMR-generated reports will summarize the utilization of FMHC services (such as initial versus follow-up ratio, clinic no-shows and patient waiting time, etc.) to improve efficiencies in clinic workflow. Data will be analyzed at six-month intervals using descriptive techniques such as frequencies, percentages, mean, and median to track change in health outcomes of patients diagnosed with HTN.

### Evaluation of Integrated Community-Based Health Information System and Patient-Centered Medical Home Model

We will use the RE-AIM framework to evaluate the effectiveness of this integrated model on HTN management at the patient and community levels. RE-AIM’s five domains (Reach, Effectiveness, Adoption, Implementation, and Maintenance) have been used in various settings to evaluate the impact of both clinical and community-based interventions, including the management of disease-focused interventions (31-33). Both quantitative and qualitative data will be collected to help with the monitoring and evaluation of the program. Process and outcome indicators will encompass community-level, patient-level, and FMHC-level data, such as site audits leading to identifying and addressing HTN management gaps.

Operational Definitions

*Hypertension*: individuals ≥ 18 years with a diagnosis of HTN

*Uncontrolled HTN*: BP readings ≥ 140/90 in individuals with a diagnosis of HTN; ≥ 130/80 for patients with diabetes mellitus and HTN

*Elevated blood pressure*: single reading of ≥ 140/90 for individuals with no prior diagnosis of HTN

*Community*: Catchment population within a 1-kilometer radius of FMHC

*CBHIS participants*: Community members who enroll in FGDs and HAS

*FMHC patients*: CBHIS participants who establish care at FMHC

*FMHC non-users*: CBHIS participants who do not establish care at FMHC

### RE-AIM Domains and Key Questions

A mixed methods approach will be used to measure outcomes related to HTN management at FMHC. Outcomes will be assessed through annual HAS data, FGDs, EMR reports, and financial data. Key questions under each dimension of RE-AIM will be assessed periodically as mentioned in Table 1, which are as follows:

*Reach*

a. What is the extent of the community’s participation in CBHIS?
b. What is the utilization of FMHC services by CBHIS participants?
c. What are the barriers to accessing care at FMHC?

*Effectiveness*

a. What is the impact of the PCMH-CBHIS model on the management of HTN amongst FMHC patients?
b. What proportion of patients with HTN received comprehensive care at FMHC?
c. What are the barriers to HTN control amongst CBHIS participants registered at FMHC?

*Adoption*

a. What is the CHWs’ capacity for ongoing community engagement activities with CBHIS participants?
b. What is the compliance of the FMHC clinical team to HTN management guidelines?

*Implementation*

a. What is the fidelity of CHWs to follow project-specific SOPs?
b. What is the fidelity of the FMHC clinical team to follow HTN guidelines?

*Maintenance*

a. What is the financial impact of implementing the PCMH-CBHIS model of care for patients with HTN visiting FMHC?

Table 1 provides key questions, outcome measures, frequency of measurement, and data sources of the study under each domain of RE-AIM.

**Table 1.**
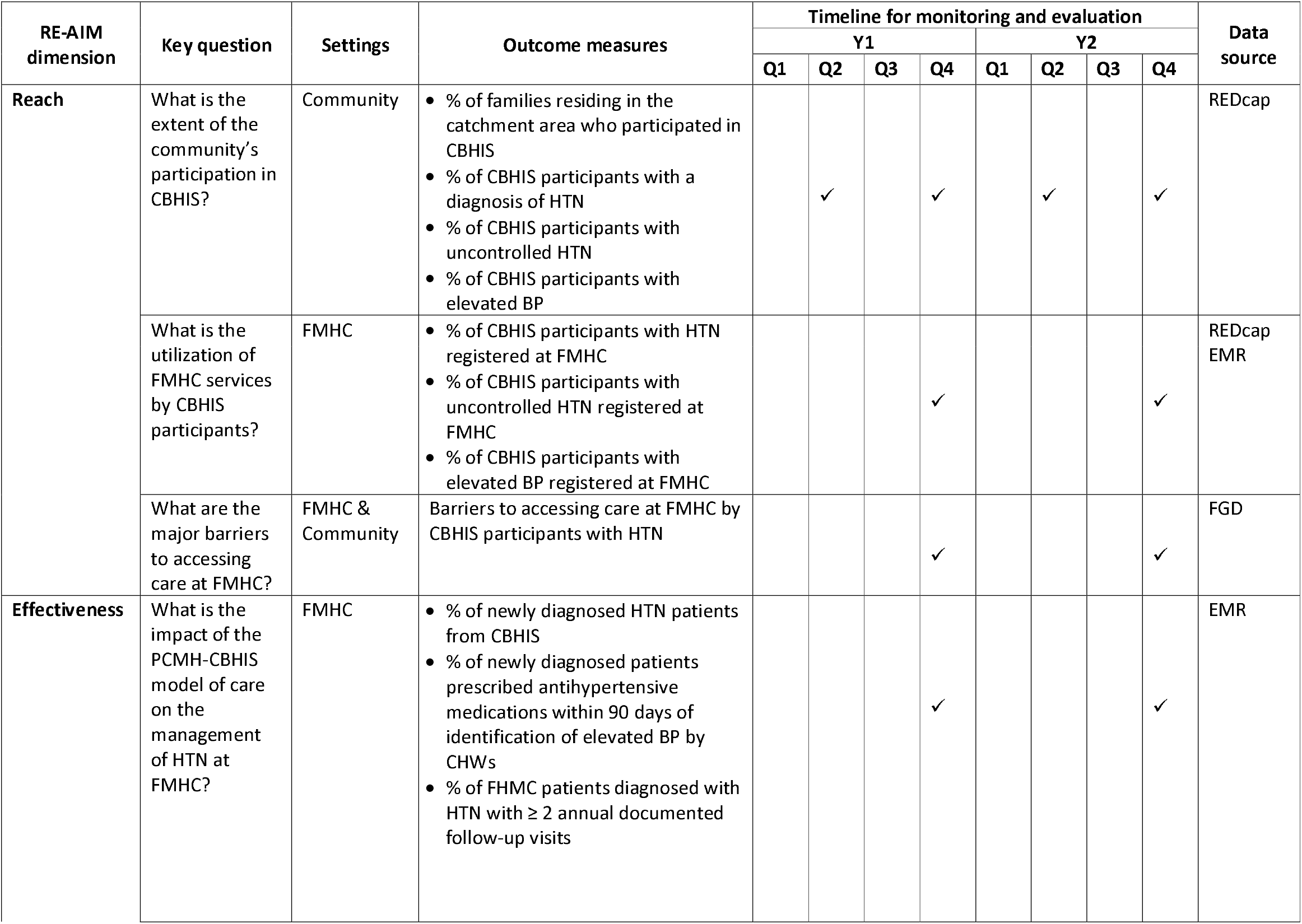
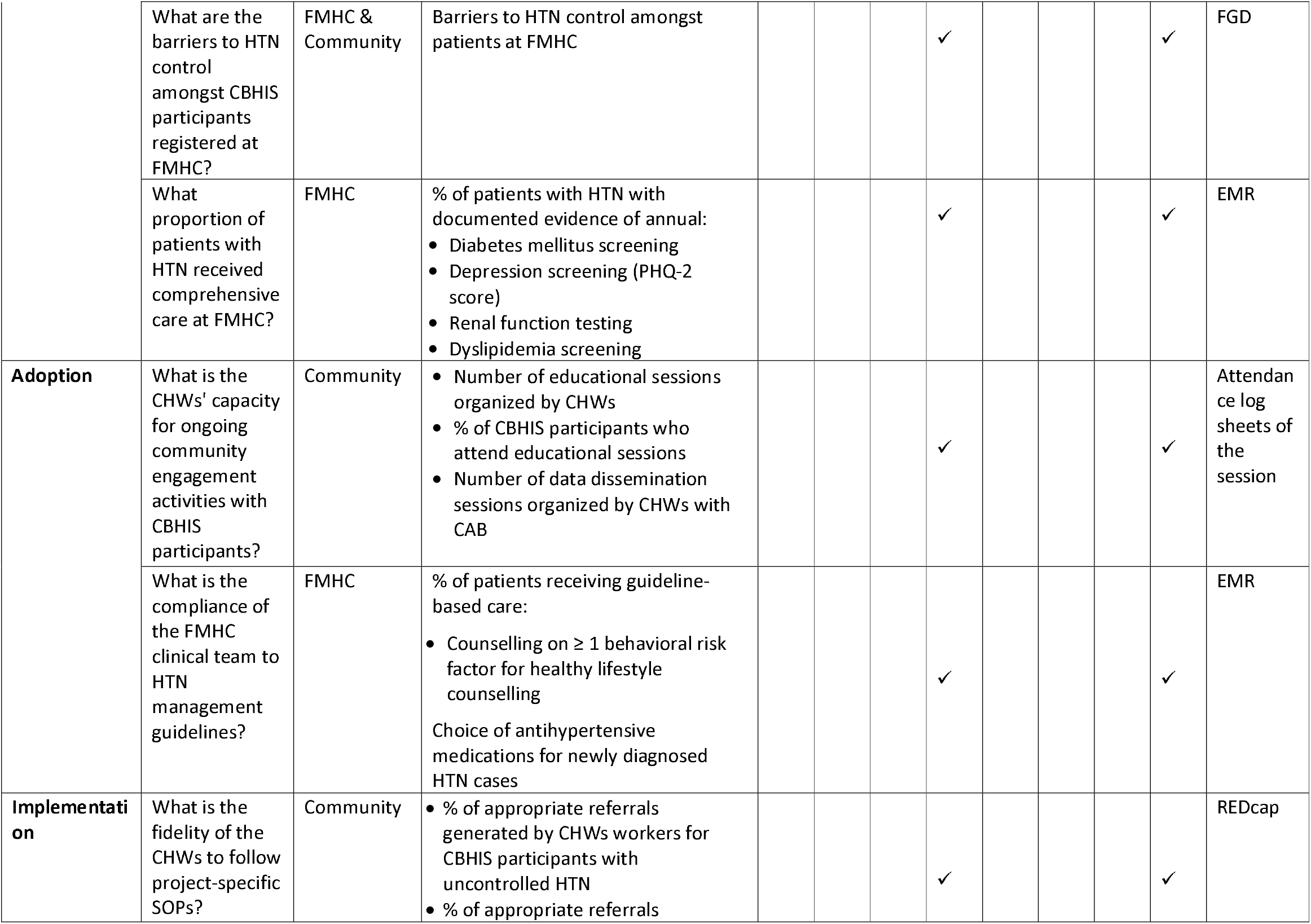
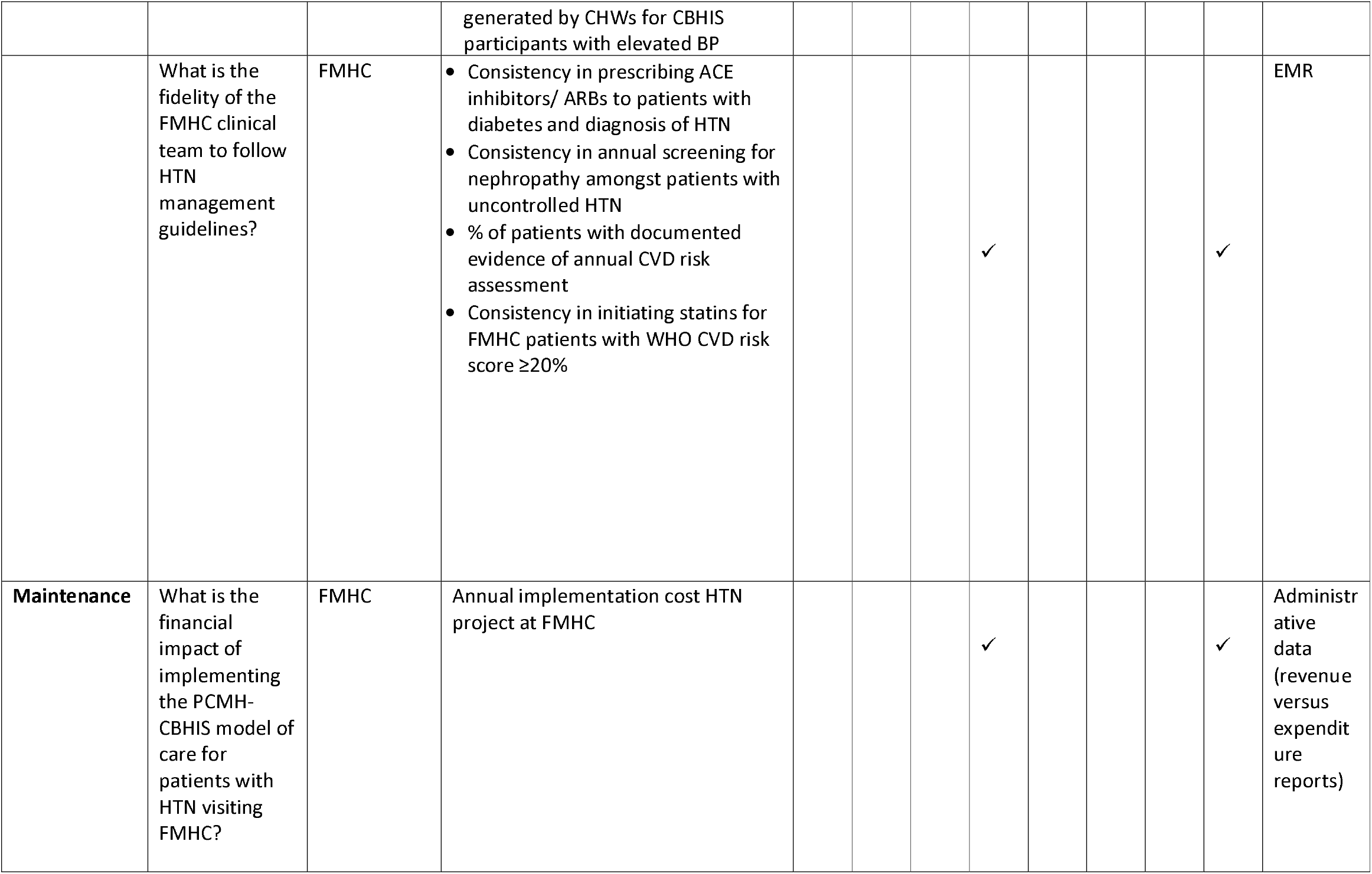
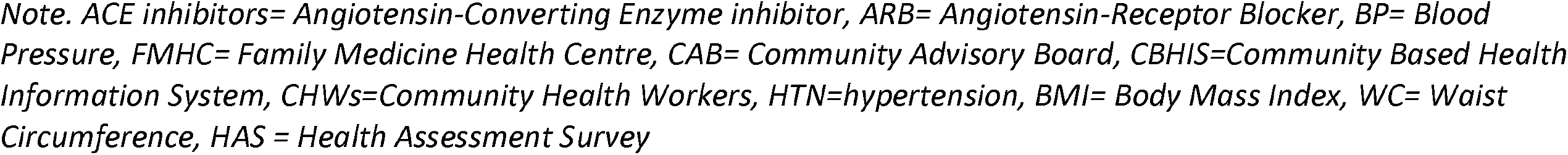
Process and Evaluation Indicators

| RE-AIM dimension | Key question | Settings | Outcome measures | Timeline for monitoring and evaluation |  |  |  |  |  |  |  | Data source |
| --- | --- | --- | --- | --- | --- | --- | --- | --- | --- | --- | --- | --- |
|  |  |  |  | Y1 |  |  |  | Y2 |  |  |  |  |
|  |  |  |  | Q1 | Q2 | Q3 | Q4 | Q1 | Q2 | Q3 | Q4 |  |
| Reach | What is the extent of the community’s participation in CBHIS? | Community | • % of families residing in the catchment area who participated in CBHIS<br>• % of CBHIS participants with a diagnosis of HTN<br>• % of CBHIS participants with uncontrolled HTN<br>• % of CBHIS participants with elevated BP |  | ✓ |  | ✓ |  | ✓ |  | ✓ | REDCap |
|  | What is the utilization of FMHC services by CBHIS participants? | FMHC | • % of CBHIS participants with HTN registered at FMHC<br>• % of CBHIS participants with uncontrolled HTN registered at FMHC<br>• % of CBHIS participants with elevated BP registered at FMHC |  |  |  | ✓ |  |  |  | ✓ | REDCap<br>EMR |
|  | What are the major barriers to accessing care at FMHC? | FMHC & Community | Barriers to accessing care at FMHC by CBHIS participants with HTN |  |  |  | ✓ |  |  |  | ✓ | FGD |
| Effectiveness | What is the impact of the PCMH-CBHIS model of care on the management of HTN at FMHC? | FMHC | • % of newly diagnosed HTN patients from CBHIS<br>• % of newly diagnosed patients prescribed antihypertensive medications within 90 days of identification of elevated BP by CHWs<br>• % of FHMC patients diagnosed with HTN with ≥ 2 annual documented follow-up visits |  |  |  | ✓ |  |  |  | ✓ | EMR |
|  | What are the barriers to HTN control amongst CBHIS participants registered at FMHC? | FMHC & Community | Barriers to HTN control amongst patients at FMHC |  |  |  | ✓ |  |  |  | ✓ | FGD |
|  | What proportion of patients with HTN received comprehensive care at FMHC? | FMHC | % of patients with HTN with documented evidence of annual:<br>• Diabetes mellitus screening<br>• Depression screening (PHQ-2 score)<br>• Renal function testing<br>• Dyslipidemia screening |  |  |  | ✓ |  |  |  | ✓ | EMR |
| **Adoption** | What is the CHWs' capacity for ongoing community engagement activities with CBHIS participants? | Community | • Number of educational sessions organized by CHWs<br>• % of CBHIS participants who attend educational sessions<br>• Number of data dissemination sessions organized by CHWs with CAB |  |  |  | ✓ |  |  |  | ✓ | Attendance log sheets of the session |
|  | What is the compliance of the FMHC clinical team to HTN management guidelines? | FMHC | % of patients receiving guideline-based care:<br>• Counselling on $\geq 1$ behavioral risk factor for healthy lifestyle counselling<br>Choice of antihypertensive medications for newly diagnosed HTN cases |  |  |  | ✓ |  |  |  | ✓ | EMR |
| **Implementation** | What is the fidelity of the CHWs to follow project-specific SOPs? | Community | • % of appropriate referrals generated by CHWs workers for CBHIS participants with uncontrolled HTN<br>• % of appropriate referrals<br>generated by CHWs for CBHIS participants with elevated BP |  |  |  | ✓ |  |  |  | ✓ | REDCap |
|  | What is the fidelity of the FMHC clinical team to follow HTN management guidelines? | FMHC | • Consistency in prescribing ACE inhibitors/ ARBs to patients with diabetes and diagnosis of HTN<br>• Consistency in annual screening for nephropathy amongst patients with uncontrolled HTN<br>• % of patients with documented evidence of annual CVD risk assessment<br>• Consistency in initiating statins for FMHC patients with WHO CVD risk score $\geq 20\%$ |  |  |  | ✓ |  |  |  | ✓ | EMR |
| **Maintenance** | What is the financial impact of implementing the PCMH-CBHIS model of care for patients with HTN visiting FMHC? | FMHC | Annual implementation cost HTN project at FMHC |  |  |  | ✓ |  |  |  | ✓ | Administrative data (revenue versus expenditure reports) |

*Note. ACE inhibitors= Angiotensin-Converting Enzyme inhibitor, ARB= Angiotensin-Receptor Blocker, BP= Blood Pressure, FMHC= Family Medicine Health Centre, CAB= Community Advisory Board, CBHIS=Community Based Health Information System, CHWs=Community Health Workers, HTN=hypertension, BMI= Body Mass Index, WC= Waist Circumference, HAS = Health Assessment Survey*

### Patient and public involvement

Patient and the public were not involved in the planning of this study.

## CONCLUSION

Our PCMH-CBHIS integrated model of care at FMHC uses a community-to-clinic approach to identify, manage, and promote self-efficacy in patients with HTN. It demonstrates a well-coordinated effort between community stakeholders, low-resource health care staff (CHWs), and the clinical team to improve the system’s performance in improving health at the individual and population levels. Existing literature provides support for the PCMH model to enhance clinical outcomes and reduce healthcare expenditures for patients with chronic illnesses (34-36). Learning from landmark studies such as the COBRA trial (37), our model builds on community engagement through CAB and CHWs since inception, inculcating a sense of ownership and empowerment that ensure sustainability (38, 39). In addition, ongoing community engagement through surveys and FGDs will allow us to explore further avenues for optimizing care at the individual, family, and community levels.

The proposed model can proactively guide changes in the health outcomes of patients with HTN. By generating evidence on the effectiveness and addressing health system-related barriers, this model will act as a proof-of-concept for strengthening the management of NCDs in primary care settings across Pakistan and other developing countries with fee-for-service health models. While we will use HTN as a prototype condition to measure health outcomes, the integrated PCMH-CBHIS model could be used to assess the efficacy of primary-care interventions for other chronic and acute health conditions.

## Data Availability

There is no data referred in this manuscript

## ETHICAL CONSIDERATIONS

Ethical approval was obtained from the Ethics Review Committee at The Aga Khan University (2022-6723-20985). Written informed consent to participate will be obtained from individual FGD participants. Family informed consent will be obtained from the family head for all family members willing to participate in HAS. They will also be informed of the voluntary nature of participation. A copy of the signed consent form shall be provided to FGD and HAS participants for record keeping. The privacy of participants will always be ensured. Time and place for FGDs and HAS will be selected within the community, in consideration of cultural norms. To ensure anonymity during FGDs no real names or directly identifying information will be reported. Additionally, FGD participants will be informed about the audio recording of the conversation. CHWs will be trained to identify, and link HAS participants with elevated blood pressure readings with FMHC clinical team for immediate care. All participants will be provided with the name, telephone number, and email address of the principal investigator.

## DISSEMINATION PLAN

The main target audience for this study includes policymakers, program implementers, health workers, researchers, and non-governmental organizations working to investigate and improve health system-related challenges in low- and middle-income countries. Moreover, the findings from this study will help improve the early detection of HTN for populations attending primary healthcare facilities. While we will use HTN to measure health outcomes, any interested group can use evidence to assess the effectiveness of the proposed model for other chronic and acute health conditions.

## ACKNOWLEDGMENTS

The authors thank Provost Carl Amrhein and Dean Adil Haider for their ongoing support.

## AUTHOR CONTRIBUTIONS

UK, JWR and SV conceived the project. UK, SS, AQ and YN planned formal and informal meetings with the community. All authors contributed towards the design of focus group discussions, household assessment survey, monitoring, and evaluation plan. UK, SS, AQ and YN led the implementation, monitoring and evaluation of the focus group discussions and household assessment survey. UK, SS and YN have contributed to the development of the electronic medical record. AM, SS, AQ and MN wrote the first draft of the manuscript. UK and SS revised the manuscript draft. All authors contributed to the refinement of the study protocol and approved the final manuscript.

